# The interplay between subcritical fluctuations and import: understanding COVID-19 epidemiology dynamics

**DOI:** 10.1101/2020.12.25.20248840

**Authors:** Nico Stollenwerk, Joseba Bidaurrazaga Van-Dierdonck, Javier Mar, Irati Eguiguren Arrizabalaga, Nicole Cusimano, Damián Knopoff, Vizda Anam, Maíra Aguiar

## Abstract

The effective reproduction ratio *r*(*t*) of an epidemic, defined as the average number of secondary infected cases per infectious case in a population in the current state, including both susceptible and non-susceptible hosts, controls the transition between a subcritical threshold regime (*r*(*t*) < 1) and a supercritical threshold regime (*r*(*t*) > 1). While in subcritical regimes, an index infected case will cause an outbreak that will die out sooner or later, with large fluctuations observed when approaching the epidemic threshold, the supercritical regimes leads to an exponential growths of infection.

The super- or subcritical regime of an outbreak is often not distinguished when close to the epidemic threshold, but its behaviour is of major importance to understand the course of an epidemic and public health management of disease control. In a subcritical parameter regime undetected infection, here called “imported case” or import, i.e. a susceptible individual becoming infected from outside the study area e.g., can either spark recurrent isolated outbreaks or keep the ongoing levels of infection, but cannot cause an exponential growths of infection. However, when the community transmission becomes supercritical, any index case or few “imported cases” will lead the epidemic to an exponential growths of infections, hence being distinguished from the subcritical dynamics by a critical epidemic threshold in which large fluctuations occur in stochastic versions of the considered processes.

As a continuation of the COVID-19 Basque Modeling Task Force, we now investigate the role of critical fluctuations and import in basic Susceptible-Infected-Susceptible (SIS) and Susceptible-Infected-Recovered (SIR) epidemiological models on disease spreading dynamics. Without loss of generality, these simple models can be treated analytically and, when considering the mean field approximation of more complex underlying stochastic and eventually spatially extended or generalized network processes, results can be applied to more complex models used to describe the COVID-19 epidemics. In this paper, we explore possible features of the course of an epidemic, showing that the subcritical regime can explain the dynamic behaviour of COVID-19 spreading in the Basque Country, with this theory supported by empirical data data.

## 1 Introduction

The SHARUCD modeling framework developed within the Basque Modeling Task Force (BMTF) to assist the Basque Health managers and the Basque Government during the COVID-19 responses is an extension of the basic epidemiological Susceptible-Infected-Recovered (SIR-type) models, and was able to describe the COVID-19 epidemic in terms of disease spreading and control, providing projections on the national health systems necessities during the first wave of the pandemic. The model was then refined to describe the disease transmission during the country lockdown [1, 2, 4] and is used, up to date, to monitor disease dynamics after social distancing measures started to be lifted (from May 4 - phase 0 and from May 11-phase 1 towards the “new normality”) [5].

The lifting of the lockdown in summer 2020 led to an increase of the infection rate with growth factors and momentary reproduction ratio hovering around the epidemic threshold (of decrease to extinction versus exponential growths). An import factor was included in the model dynamics after the full lockdown lifting in July. Although this factor was not important during the exponential growths phase in March, 2020, undetected imported infections play a major role during the stochastic phase pre/post exponential phase, where only small number of infections are detected. It refers to infected individuals (most likely asymptomatic) coming from outside the studied population (either an infected foreigner visiting the region or an infected Basque returning to the country) that are not detected by the current testing strategy. Note that when the community transmission is under control (social distancing, masks and hygienic measures), the import factor does not contribute significantly to the epidemic, only starting isolated outbreaks (variable sizes depending on the momentary infection rate), but not driving the current epidemic into a new exponential growths phase.

The effective reproduction ratio *r*(*t*) threshold of an epidemic controls the transition between a subcritical regime (below the epidemic threshold) and a supercritical regime (above the epidemic threshold). While the super- or sub-critical regime of an outbreak is often not distinguished, its behaviour is of major importance to understand the course of an epidemic since in subcritical regimes, an index infected case will cause an outbreak that will die out sooner or later, with large fluctuations observed when approaching the epidemic threshold, while the supercritical regimes leads to an exponential growths of infection. Although nowadays *r*(*t*) is largely used in the public media, an eventually better measure of decline or increase of an epidemic is the exponential growth rate *λ*(*t*), being negative below threshold or positive above threshold, see [2] for a more detailed discussion.

As a continuation of the COVID-19 Basque Modeling Task Force (BMTF), we now investigate the interplay of large fluctuations on disease spreading dynamics [6] and import in sub-threshold community spreading regime, characterized by increased mobility but still under restrictions like wearing masks and social distancing. Without loss of generality, we first describe the interplay between critical fluctuations and import in simple models of Susceptible-Infected-Susceptible (SIS) and Susceptible-Infected-Recovered (SIR) types, taking into account previous results on directed percolation [10, 9] and dynamical percolation [7, 8], to characterize the dynamics of large fluctuations during an epidemic. As this same dynamical behaviour is also observed in more complex models, the results are then generalized to the current SHARUCD modeling framework to explore possible features of the course of the COVID-19 epidemic in the Basque Country. The analysis is mainly based on the development of severe cases, i.e. hospitalizations, intensive care unit (ICU) admissions and deceased cases, while the variation of total number of positive COVID-19 cases is largely due to changes in testing capacities.

Sub-threshold community spreading leads to a decline in number of disease cases, but a small import factor is able to ignite isolated outbreaks, occasionally of large sizes, which can lead to a stationary number of new cases in mean field approximation, even when large fluctuations around it are observed. These fluctuations are larger when closer to the epidemic threshold and such increased numbers of cases are treated by many people as “second waves” and compared with the first exponential phase in March and April, 2020, where community spreading was definitely well above the critical threshold. Here, we reserve this term to a new exponential growths phase expected in a supercritical community spreading regime, where the number of cases are significantly larger than the actual cases referring to subcritical dynamics with import. The 20 days predictions provided to the Basque Health managers and the Basque Government were obtained by assuming that the community transmission is under control, below the threshold, not increasing to a new exponential phase at the time. That has being proven to hold since August 2020 well into late October 2020.

With real data supporting the theory, in this paper we show that the sub-critical regime can explain the dynamic behaviour of COVID-19 epidemic in the Basque country, at least until end of October and beginning of November 2020, when new lockdown measures where implemented. This result might explain the COVID-19 epidemic behaviour in many other European regions as well during the late summer period and autumn. The idea that this “second wave” was under control as the new lockdown measures contribute to decrease the community transmission ever more, eliminating larger subcritical fluctuations, brings up the discussion of an eventually “third wave” while the control measures are again relaxed. Here we also discuss the current epidemiological scenario of COVID-19 in the Basque Country, showing that although it is still early to know to which lower level the disease transmission will develop, the current model continues to predict stationarity for all variables. Community transmission might have reach the supercritical regime during winter due to well known seasonal effect of other respiratory diseases if new lockdown measures were not taken.

The presently updated version of this manuscript as of March 2021 add to the original version of December 2020 a short discussion on reproduction ratio r(t) hovering around the epidemic threshold, even though the community spreading is constant below threshold *β* ≲ *β*_*c*_, due to the fluctuations of isolated outbreaks after imported index cases. Also added as appendix is an inspection of COVID-19 data of some European countries between introduction in Spring 2020 until late autumn 2020.

## 2 The role of import in epidemiological systems close to the epidemic threshold

### 2.1 A general SIR epidemiological model with critical threshold and import

Basic epidemiological systems which capture many important issues of disease spreading dynamics can be phrased as SIR-type models, with a population of *N* individuals divided into susceptible individuals *S* to an infectious disease under consideration, infected individuals *I* and recovered individuals *R* which are either immune against the disease during their entire life, like in childhood diseases, or return to become susceptible again, often due to mutations of the pathogens, or gradual waning immunity of the hosts.

In the studied population *N*, besides considering that infected individuals transmit the disease to a susceptible individual, which then turn to be infected as well, we also consider an import factor which refers to the possibility of susceptible individuals inside the studied population becoming infected by an undetected infection chain started outside the studied population, i.e. infected individuals (most likely a mobile asymptomatic infected individual), either a foreigner visiting the region or a local returning to the country without being detected by the current testing strategy.

The basic model for the expected mean number of susceptible, infected and recovered individuals reads as follows

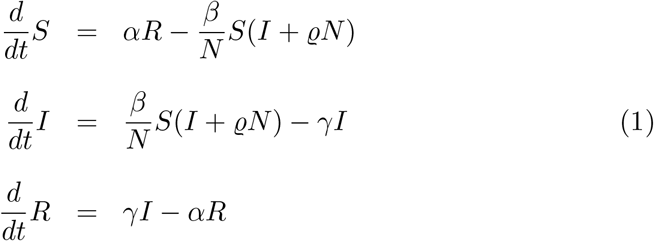

with transition probabilities per time unit of infection rate *β*, recovery rate *γ* and waning immunity rate *α* and as a less well studied feature the import ratio *ϱ*.

For our COVID-19 modeling framework we consider in the infection class *I* a division into mild/asymptomatic individuals *A* and severe/hospitalized individuals *H*, as well as disease induced death *D* and intensive care unit admitted patients *U*. These classes are recorded as incidences of cumulative cases *C*, giving a SHARUCD modelling framework to be compared with the empirical data at hand. For complete descriptions of this modelling framework and its application to COVID-19 epidemic in the Basque Country, see [1, 2, 4, 5].

The above given dynamical system, Eq. (1), describes the mean field approximation of an underlying stochastic process given by the dynamics of probabilities *p*(*S, I, R*). Since *S*(*t*) + *I*(*t*) + *R*(*t*) = *N* we only need to consider *p*(*S, I*), and from this we can conclude *R* = *N* − *S* − *I*. The process is given by the following governing equation system,

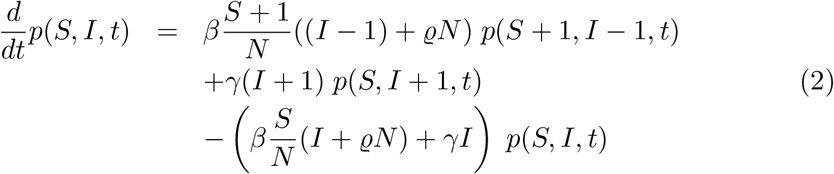

called master equation in chemistry and physics and known, in mathematics, as state discrete and time continuous Markov process, here for long term immunity *α* → 0, described in more detail e.g. in [6] and its references to earlier literature.

With an appropriate numerical scheme, the so-called Gillespie algorithm also known as minimal process algorithm, we can immediately investigate the qualitative behaviour of such an epidemiological system as shown in Fig. 1.

**Figure 1:**
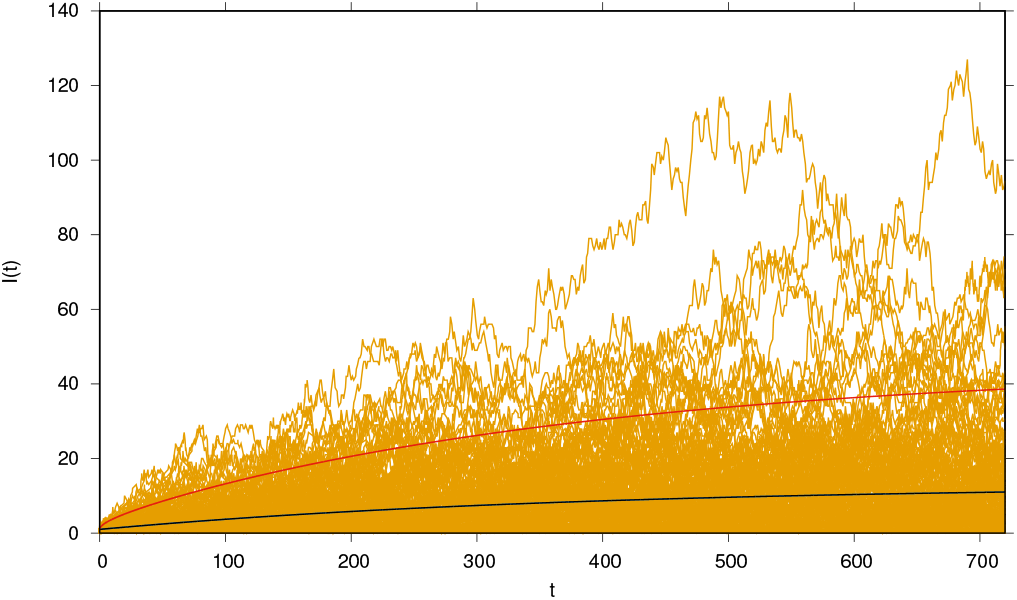
Ensemble of realizations of the number of infected with very wide excursions into large outbreak regions. Parameters are in the subcritical regime but close to the epidemiological threshold. Mean dynamics and variance dynamics give the quantification of the fluctuations. Black line gives mean dynamics of ⟨I⟩, and red line gives two standard deviations on top of the mean, obtained from the variance dynamics of V (t) ≔ ⟨I^2^⟩ − ⟨I⟩^2^. The large excursions of some of the stochastic realizations indicate long tail distributions rather than Gaussian distributions around the mean values (remembering that µ ± 2 · σ gives roughly 95% of the Gaussian distribution). The essential parameters are β = 0.95 γ, close to the epidemiological threshold of β = β_c_ = γ and small import ϱ = e^−15^.

We start the simulations each time with one infected individual which acts as a first index case for a small or larger outbreak, dieing out after some time due to the infection rate being smaller than the recovery rate, i.e. the epidemiological system is subcritical.

Even though the initial outbreak from this first index case terminates earlier or later, due to the small probability to generate new index cases, new outbreaks are generated by index cases from import *ϱ*, often dieing out quickly but occasionally causing big excursions into avalanches of large numbers of infected, since the community spreading *β* is just below the threshold of negative growth, i.e. *ε* ≔ *β* − *β*_*c*_ here with *β*_*c*_ = *γ* is negative *ε* < 0, versus positive growth where *ε* > 0.

This import factor was included in the SHARUCD modeling framework right from the beginning of the pandemic, in March 2020, but the initially available data could not give any information at the starting of the epidemic, when usually undetected imported infections play a major role on the development of the outbreak. Although this factor was no longer important during the exponential growth phase, it has become essential again during the lockdown-lifting phase in summer/autumn 2020, to understand the dynamics in combination with *ε* = *β* − *γ* ≲ 0, the sub-threshold community spreading, whereas an already new “second wave” would lead with *ε* ≫ 0 to a new exponential growth, which was eventually to be expected in winter due to seasonality observed in respiratory diseases, naturally increasing the community spreading *β*(*t*).

### 2.2 The SIS model with import and the SIR general epidemic process (GEP): well studied limiting cases around criticality

From the above described SIR model with import, Eq. (1), and its stochastic counterpart, Eq. (2), some interesting simplified models can be derived as special cases in certain dynamical regimes, which have been studied extensively in the literature over decades.

From the SIR model with import the SIS model with import can be derived by neglecting the recovered class or assuming the transition from *R* to *S* as infinite *α* → ∞ and with conserved population size *N* = *S*(*t*) + *I*(*t*), hence *S* = *N* − *I*, given by its mean field deterministic version

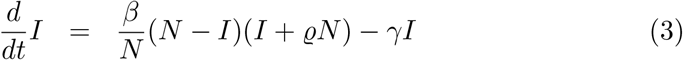

and from this the corresponding stochastic process is easily elaborated as well [11].

The SIS epidemiological model even without import is considered as a paradigmatic system for a whole universality class in its dynamical behaviour around the critical threshold, directed percolation [10], and the role of import as a conjugated field around the critical behaviour was more recently investigated in great detail in [9]. The behaviour of systems around critical thresholds has been investigated in equilibrium thermodynamic systems like the Ising spin system since a century and more recently the notions have been transfered to non-equilibrium critical threshold behaviour, where the SIS model in its spatially extended version is a prime example. The conjugated field in the Ising model is an external magnetic field giving preference to one direction of magnetization of the system over the other.

The spatially extended SIS system in its simplest form is given by the stochastic process with probabilities of configurations considering one or non infected *I*_*i*_ at each site *i* of any network or spatial grid

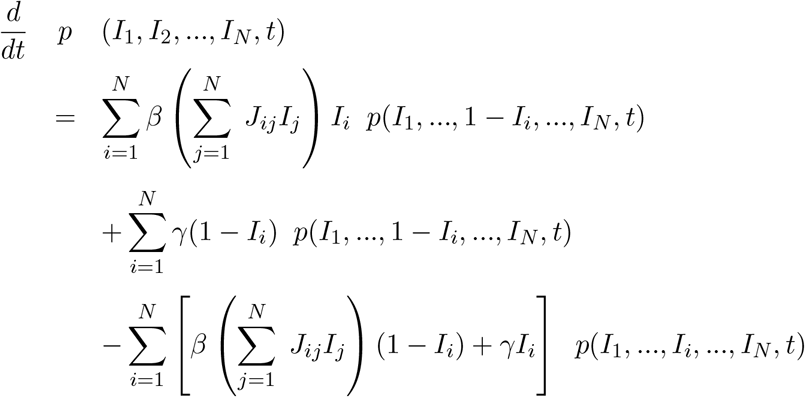

again with infection rate *β* and recovery rate *γ* and an adjacency matrix *J*_*ij*_ specifying which network site *j* is neighbor to *i* and can be a link of infection, see [6] for detailed description and further considerations of such systems. Any more extended SIR system in spatially extended stochastic settings can be given respectively, see e.g. [12].

Such systems show around the critical threshold a power law behaviour with universal power law exponents numerically equal for whole classes of models with only few common features like dimensionality, i.e. connectivity in generalized networks, and existence e.g. of absorbing states. In the case of the universality class of directed percolation an absorbing state is needed, here in the SIS system characterized by zero number of infected, from which no new infection can be generated, see for a general discussion e.g. [9]. When the connectivity of a spatial system or network is large enough the exponents reach their mean field values which correspond to the dynamic behaviour of the above described non-spatial systems. This threshold of connectivity can be reached either in regular spatial systems above a certain dimensionality, the critical dimension, in the case of directed percolation e.g. four dimensional systems, or by spreading with jump distributions with diverging variance, i.e. Lévy flights in superdiffusive spreading systems. On superdiffusive spreading in the framework of the above mentioned Eq. (4) see e.g [13] and further references there, and for the role of such superdiffusive connectivity in the transition between spatial two-dimensional critical behaviour to mean field behaviour see as a very recent publication [8].

Here we only consider the mean field versions of such epidemiological systems, since most of the time a high mixing of populations drive most real world epidemiological systems towards the mean field behaviour, so that this is always the starting point of any analysis, but we always consider the mean field dynamics as limiting case of much wider dynamics in any connectivity stochastic system, where around critical thresholds only few system properties are important and any microscopic details become unimportant under the large fluctuations observed around threshold. Further we consider the SIS dynamic as an initial studying field since here many aspects at least in its mean field behaviour can be studies analytically, developing the methodological terminology in a rigorous and controlled way, and then relying in more complex systems increasingly on numerical simulations and eventually approximations as in the case of critical exponents derived from field theoretical analyses, known in theoretical physics for many decades and increasingly being applied also to non-equilibrium phase transitions.

In [10] the notion of stochastic processes like the SIS dynamics being in one universality class with the same critical exponents for all models in this class was established, and the exponents were characterized as being the ones of “directed percolation”, i.e. percolation in spatial systems with certain dimensionality plus one dimension of time, which has one direction but does not allow infection back in time. Its mean field dynamics is given by an equation equivalent to Eq. (3) with *ϱ* = 0. Small perturbations and scaling around criticality can be investigated by including the import *ϱ* > 0 as a conjugated field, like an external magnetic field in the Ising spin universality class, as has been extensively investigated in [9].

On the other hand another universality class has been attributed to the SIR epidemiological system spreading from an initial source into a large or asymptotically infinite system of susceptible with critical exponents equal to the ones observed in ordinary percolation for the remaining cluster of recovered after a wave of infection spreading out, the universality class coined as “dynamical percolation”, see [7]. While this universality class has been studied in great detail also for super-diffusive spreading giving an interpolation of dynamic behaviour and hence critical exponents between purely local and mean field behaviour, see e.g. [8] and many related references there, no consideration of the role of import has come yet to light for the case of dynamical percolation, so that we study here with the notions of the SIS system with import also now the SIR initial spreading critical behaviour under the influence of import, in order to understand qualitatively the behaviour in more complex models suitable for the description e.g. of the spreading of COVID-19 into an initially susceptible population with not yet having reached any solid evidence of growths restriction by e.g. herd immunity which would ultimately limit the availability of “resources” in form of susceptibles.

The SIR system in its spreading phase is again a limiting case of Eq. (1), namely this “general epidemic process” (GEP) [7] follows from the SIR model via the limits of vanishing waning immunity *α* = 0 and vanishing import *ϱ* = 0 as

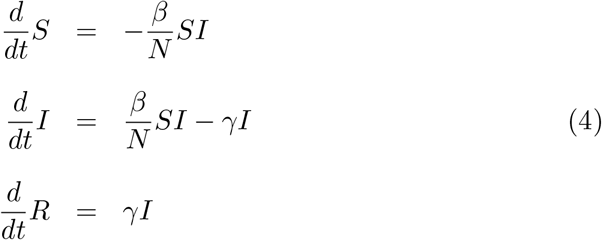

hence leaves a remaining cluster of recovered *R* after the infected spread out, which are not removed due to long lasting immunity, with a time scale *α*^*−*1^ → ∞ much longer than the infection process. Again, spatially extended stochastic versions of this GEP can be given, and its non-spatial stochastic and deterministic mean field counter parts give a good analysis when the connectivity between individuals is reasonably large.

### 2.3 The SIR model with import around the critical threshold

The stochastic realizations in Fig. 1 are obtained via the complete stochastic process described by Eq. (2), such that the susceptible individuals could still be burned out after a while, once a first index case and subsequent imports start some outbreaks.

When we assume a large enough pool of susceptibles, as is done in the initial disease spreading analysis in the GEP giving rise to criticality of “dynamical percolation” type [7], we obtain from Eq. (2) by assuming that in the initial phase of an epidemic with nearly the whole population still susceptible *S/N* ≈ 1 the dynamics for the number of infected is now given by

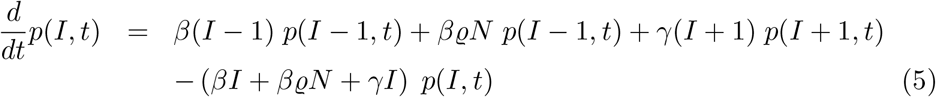

from which we can easily calculate the time dependent mean value 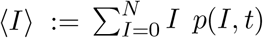 and its variance *V* (*t*) ≔ ⟨*I*^2^⟩ − ⟨*I*⟩ ^2^ analytically. The results are given in Fig. 1 for the mean ⟨*I*⟩(*t*) as black line, and from the variance *V* (*t*) we plot two standard deviation above the mean as red line. Since the distribution is highly non-Gaussian we observe the mean close to the extinction boundary of zero numbers of infected and any lower variance cannot be described by the above used method. Further the upper variance is exceeded by many stochastic realizations in yellow, indicating a long tail distribution typical for a system close to a critical threshold.

The analytic expressions are for the mean number of infected, when starting exactly with e.g. *I*(*t*_0_) = 1 infected,

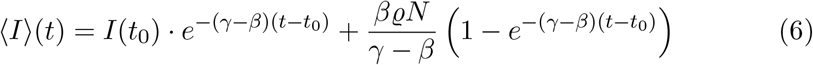

here organized for *β* < *γ*, hence controlled community spreading (but also holding in the exponential growth case *β* > *γ*), and for the variance we have

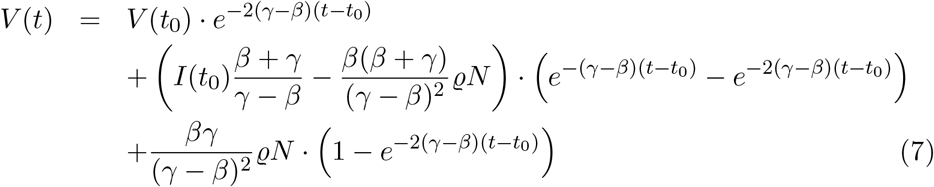

such that for subcritical behaviour *β* < *γ* both, mean value and variance, reach a finite stationary state, namely the mean value 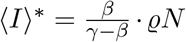 and the variance 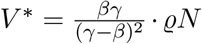. Hence no matter how large the import is, it will always reach a finite level and not explode exponentially.

We first investigate how the behaviour changes for increasing import, see Fig. 2 a). Also here the mean value and the variance reach a stationary state, i.e. they level off after an initial increase, as long as the community spreading is below the epidemiological threshold, i.e. the growth factor *ε* ≔ *β* − *γ* < 0. When the growth factor of community spreading is above the epidemiological threshold, i.e. *ε* > 0, we observe exponential growth, see Fig. 2 c), not limited by any exhaustion of susceptibles, since we assume *S/N* ≈ 1.

**Figure 2:**
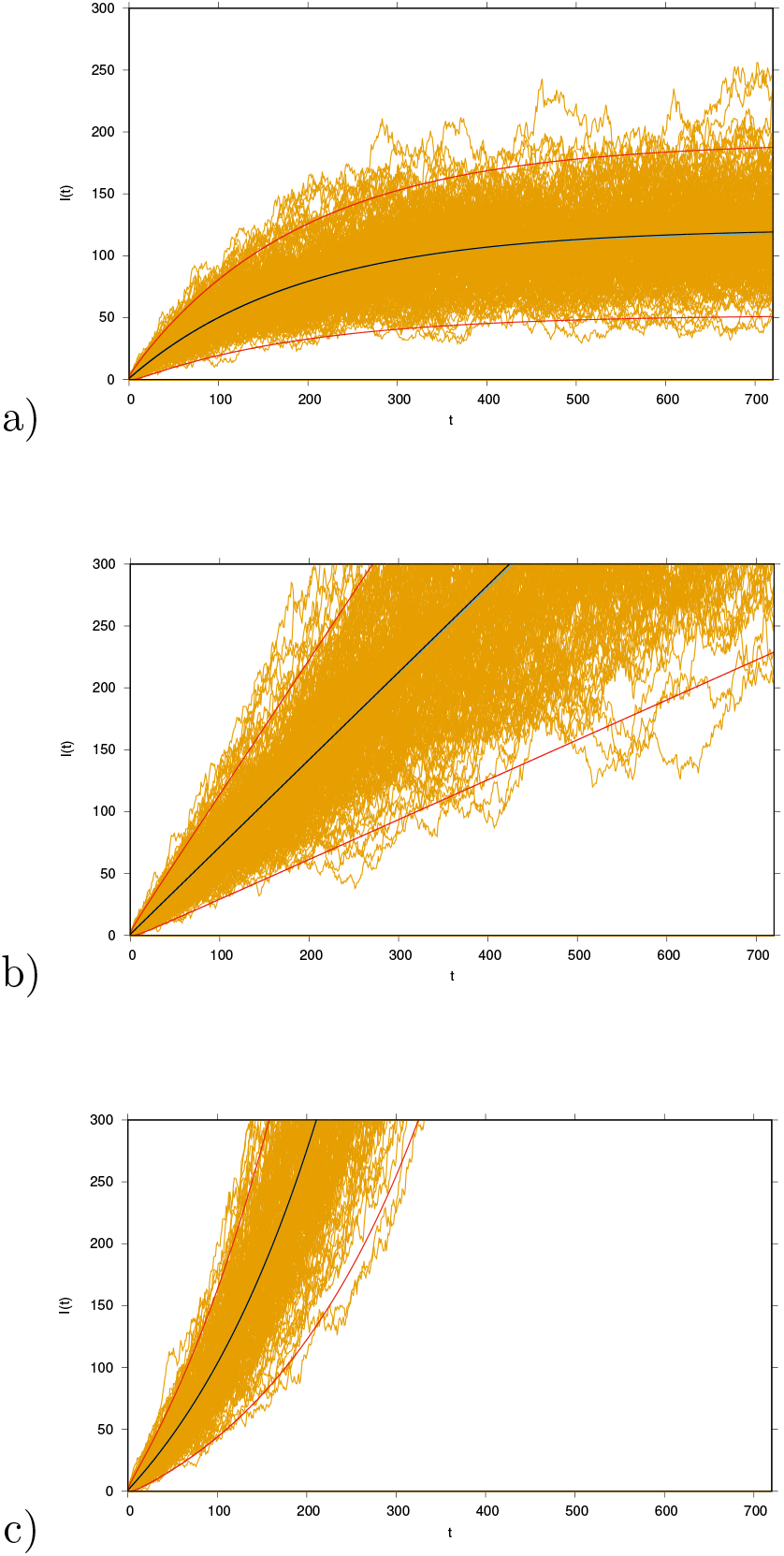
Mean dynamics and variance curves (*µ* ± 2*σ*) and stochastic realizations of SIR model with import. Increased import *ϱ* = *e*^*−*12^. Variation of *β* with a) *β* = 0.9 · *γ*, b) *β* = *β*_*c*_ = *γ* and c) *β* = 1.1 · *γ*.

Then we investigate the dynamic behaviour of the system closer to criticality under constant import, see Fig. 3. The closer we are to the critical threshold with its linear growth the longer the mean values stay close to the critical line, but then subcritically always level off to its stationary state, and above threshold always finally explode into an exponential phase.

**Figure 3:**
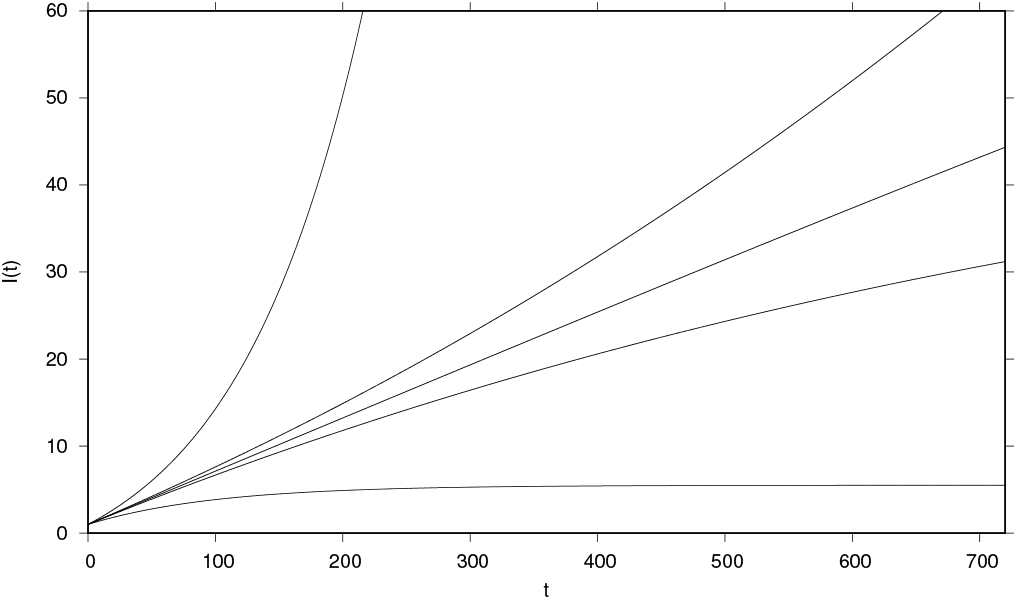
For community infectivity very close to criticality β = 0.99 · γ and β = 1.01 · γ the solutions appear initially close to the critical line of β = β_c_ = γ, but finally level off subcritically again, respectively explode into the exponential phase. Analytic mean field solution from bottom to top for β = 0.9 · γ, β = 0.99 · γ, β = β_c_ = γ, β = 1.01 · γ and β = 1.1 · γ, in all cases import ϱ = e^−15^ and N = 2 · 10^6^ and γ = 0.05 d^−1^.

Finally at the critical threshold, i.e. *β* = *β*_*c*_ = *γ*, we have a linear growth of the mean number of infected with time, analytically given by

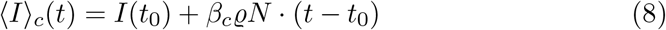

hence in terms of critical behaviour a power law growth with mean field exponent 1. Since ⟨*I*⟩ _*c*_(*t*) ∼ *t*^1^ we also can infer for the mean number of recovered ⟨*R*⟩_*c*_(*t*) ∼ *t*^2^, due to the dynamic equation 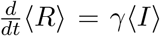, which confirms the scale-free dynamical behaviour in form of another power law, see Fig. 4, with straight lines in double logarithmic plots for ⟨*I*⟩_*c*_(*t*) and ⟨*R*⟩_*c*_(*t*), and deviations from this for values smaller and larger *ε* ≠ 0.

**Figure 4:**
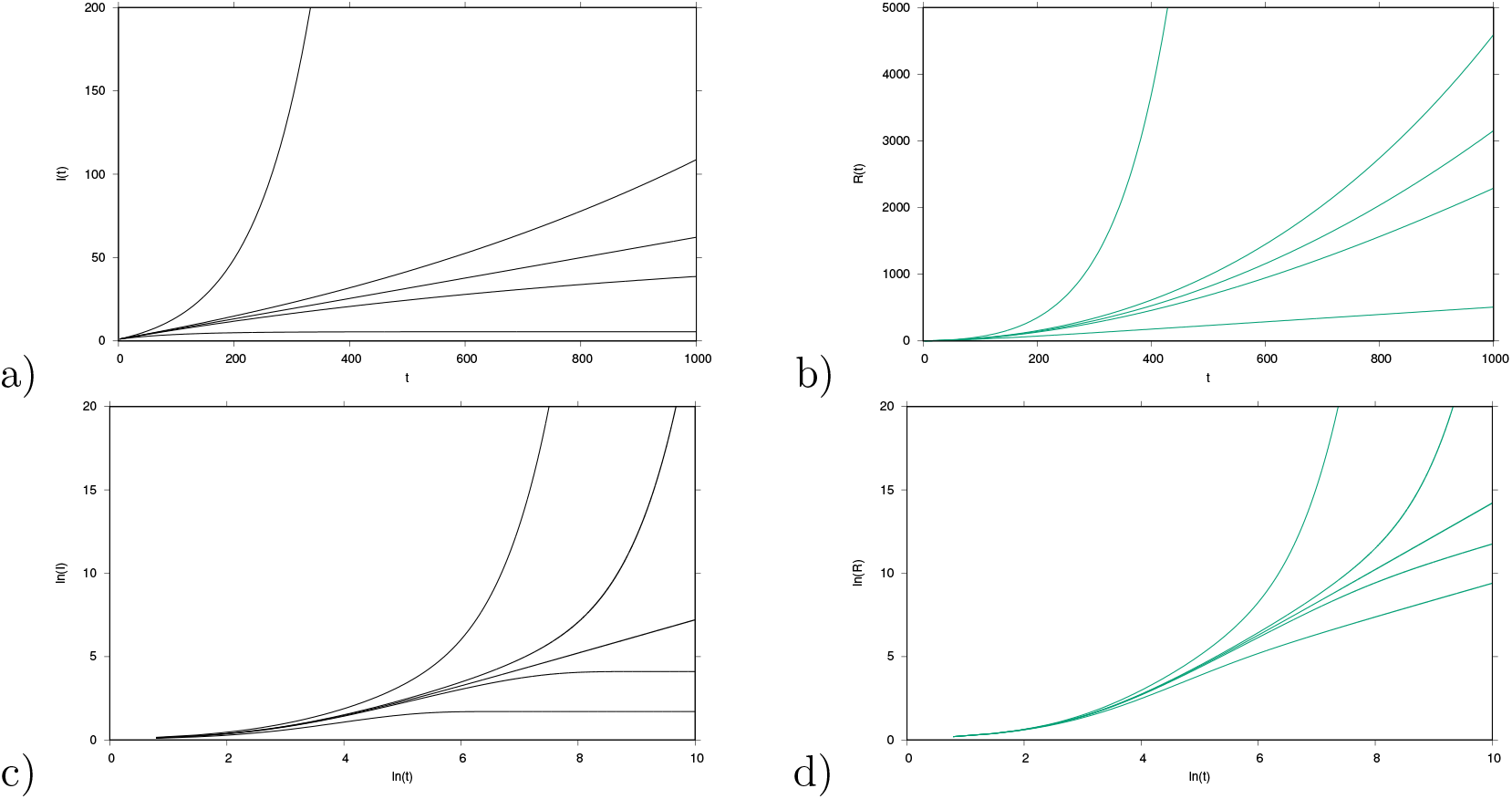
The mean field solutions for subcritical, critical and supercritical SIR models with infinite susceptible individuals in natural scale. a) Infected from bottom to top for β = 0.9 · γ, β = 0.99 · γ, β = β_c_ = γ, β = 1.01 · γ and β = 1.1 · γ, in all cases import ϱ = e^−15^ and N = 2 · 10^6^ and γ = 0.05 d^−1^. The critical curve shows ⟨I⟩ (t) ∼ t. b) Recovered for the same parameters. The critical curve shows ⟨R⟩(t) ∼ t^2^, which here in natural scale appears still as an upward bent curve. In c) and d) the mean field solutions for subcritical, critical and supercritical SIR models with infinite susceptibles in double logarithmic scale. in c) for the infected with the critical curve in straight line, showing ln⟨I⟩(t)) ∼ 1 · ln(t), and in d) for the recovered, for which the critical curve shows ln ⟨R⟩(t)) ∼ 2 · ln(t) which appears now also as straight line separating the subcritical and the supercritical regimes.

For such spreading into an environment of abundant numbers of susceptibles, already without import, one observes the asymptotic behaviour around criticality given by

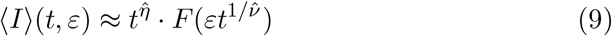

with critical power law exponents 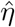 and 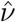, see e.g. [8] and there Eq. (15), holding for small *ε* → 0 and long times *t* → ∞ and *F* a universal scaling function. The well established values of these critical exponents in 2-dimensional spatially extended stochastic systems, respectively in mean field approximation (m.f.) are

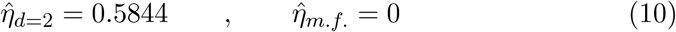

and

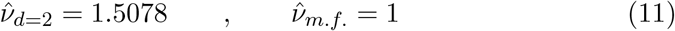

see [8], there Figs. 16 and 19. The mean field exponents can in complete agreement also be read off from our analytical expression, Eq. (6) here. For the scaling with import no such results are established, but our mean field approximation gives also here an exponent of 1. The scaling with import however has been well studied in the SIS case, and the exponents there are already in mean field approximation more non-trivial, deviating from 0 or 1 and hence this SIS case with import is a good studying field for the concepts described here, see for a good exposure [9], but has to be distinguished from the present case of relevance, the SIR system in the spreading regime of abundant numbers of susceptible individuals. A more complete discussion of the framework of scaling around criticality can be found in the appendices.

### 2.4 A single stochastic realization of the subcritical SIR model with import and estimation of its reproduction number *r*(*t*) hovering around the epidemiological threshold

We finally give an example of a single stochastic realization of infected of the subcritical SIR model with import with parameters as used in Fig. 1. This example realization we then use as data set to analyze the momentary reproduction rate *r*(*t*) as described in detail in [2], and as commonly used nowadays also in the public media to monitor the ongoing COVID-19 epidemical situation.

In Fig. 5 a) one of the realizations of the ensemble of Fig. 1 is shown and for better orientation also the mean and two standard deviation lines. The present realization is not untypical since it lies most of the time in the 95% confidence interval and does not even show the often observed large deviations seen in Fig. 1, though quite some realizations also die out quickly and eventually later bounce back due to the small import probability. When we calculate from such a realization of the subcritical process, remembering that *β* = 0.95 · *γ* < *β*_*c*_, the reproduction ratio *r*(*t*) via a sliding window, as described in detail in [2], we observe that due to the fluctuations in the numbers of infected the measured reproduction ratio hovers around the epidemiological treshold of *r*_*c*_ = 1, sometimes above and sometimes well below one, see Fig. 5 b). For graphical reasons we only show the first 180 days, but the effect is well visible throughout the entire time series from Fig. 5 a). The theoretical value for the reproduction ratio of a simple SIR model without import is *r* = *β/γ* as long as the susceptibles are abundant and not show any signs of being burned out yet. This gives the green line in Fig. 5 b) below the threshold value of *r*_*c*_ = 1.

**Figure 5:**
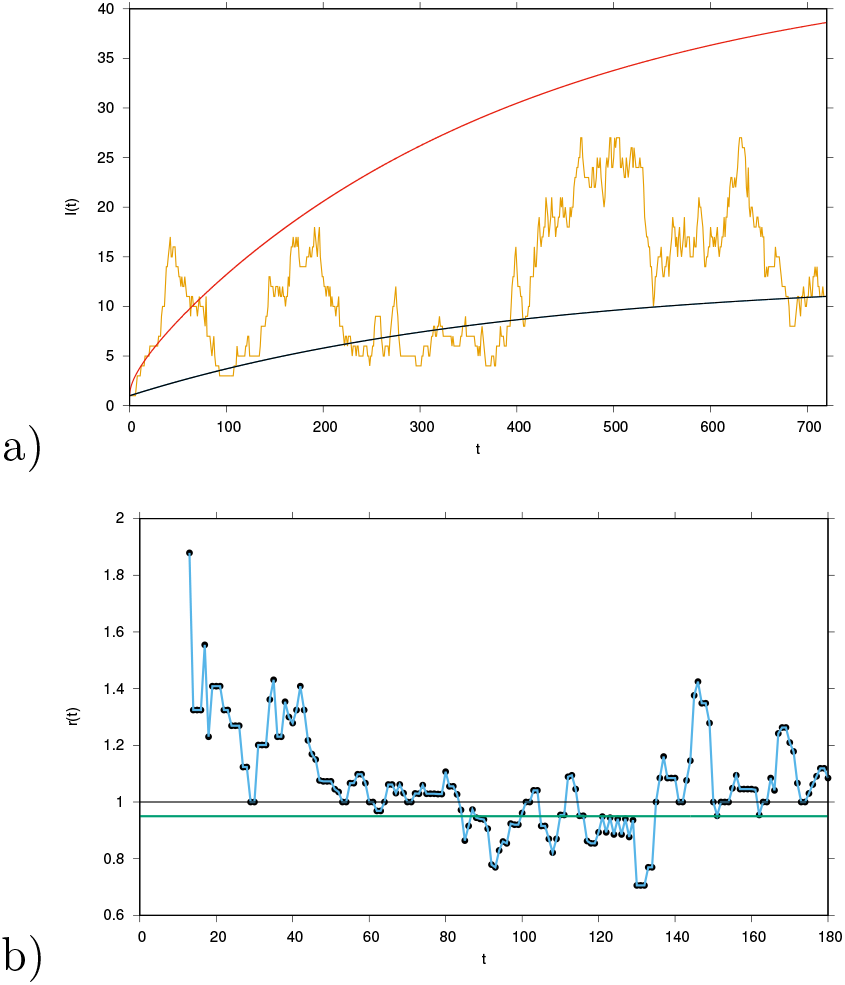
a) Single stochastic realization of the subcritical SIR model with import and constant infection rate β < β_c_ and b) from this calculated the momentary reproduction ratio r(t) hovering around the epidemiological threshold of r_c_ = 1, though the theoretical value for a simple SIR model is the fixed quantity r = β/γ = 0.95 < 1 for community spreading (green line).

The simple SIR model without import has the reproduction ratio *r* = *β/γ*. To measure *r* as indicator of the community spreading, i.e. if the epidemic is growing or declining in a given study population, one would have to detect every new imported case as index case and measure all secondary cases from every index case, especially also the isolated index cases which create none or eventually one secondary case, to obtain accurate statistics of the community spreading and its capability of sustaining transmission or not.

This is practically near to impossible in COVID-19 due to many asymptomatic cases never being detected. And any momentarily used empirical measure of the growth rate *r* tends to only account for the larger detected outbreaks and does not distinguish between new community infected cases and cases from any “import probability”. Hence they tend to overestimate the reproduction ratio *r*, as being defined as production of new cases in a given time interval devided by the new cases in a previous time interval.

Especially when not distinguishing between community infected and import infected there appear conceptual problems: from a disease free state a single infected new case gives an infinite reproduction ratio when taking the ratio of new infected over previously infected [20] as basis for any reproduction ratio calculation, i.e. 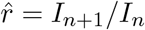.

The way forward here is to simply take the stationary solution of the SIR system with import, which has on average as many cases in the new infection period as in the previous infection period, hence the estimated reproduction rate 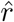 is exactly unity 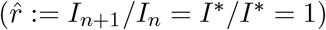. Still, the epidemiological system is subcritical in the sense discussed above of not being exponentially increasing. This shows once again the limitation of the concept of reproduction ratio, since the imported cases are eventually reproduced outside the study population, but do not give any indication of the system under study of exponentially growing or exponentially declining dynamics. Only a response consideration, i.e. stochastic excursion outside the stationary mean value and its time to relax to stationarity could eventually give a better picture of the epidemiological dynamics inside the study population.

In any case, all classical considerations of reproduction ratios are based on initial estimations of exponential growth rates of infections, and insecurities arise mainly from largely unknown next generation times [21], hence the exponential growth rates are of primary interest and often more informative than reproduction ratios [2]. Ultimately, the question of managing an epidemic is about exponential growth up to exhaustion of susceptible or exponential decline towards a stationary state.

The present use of the term “import” is quite broad and means any probable infection of a susceptible of the study population by unknown or unreported causes, may be by external infection, by unaccounted asymptomatic chains of infection or even long range infections not accounted for in close contact tracing considerations. Frequently, in field epidemiology of the present COVID-19 pandemic frustrations are expressed about the detection of many sparking events not linked to any known and securely detected infection chains, highlighting the need of describing such events statistically in the way done here.

From this observation follows clearly that for any epidemiological system with community spreading just below the critical threshold it is difficult to estimate the reproduction ratio since this would oscillate above and below threshold due to purely stochastical fluctuations which are most pronounced around the critical threshold. In COVID-19 we observe at the moment in many countries reproduction ratios reported just above or below *r*_*c*_ = 1 and relatively frequently crossing the threshold.

## 3 COVID-19 epidemiological dynamics in Euskadi, País Vasco: from the initial phase up to after the summer lockdown lifting

To model the dynamics of the epidemiological spreading of COVID-19 during its initially exponential growth phase and into the lockdown control phase we developed earlier a modelling framework on the basis of simple SIR-type models, refining to distinguish severe hospitalized cases from mild or even asymptomatic cases, which might also at times go unnoticed, in a SHAR-type model [4], and then adjusted to describe additionally available data on intensive care unit (ICU) admitted cases and deceased in the final framework of SHARUCD models [1, 2]. Detailed descriptions of models and their adjustments to available data from the Basque Country can be found in [5].

At the time we included formally already an “import term” as described in the SIR-type models above, but did not have sufficient information to infer its role in the dynamics, first due to the large exponential growth and later due to travel restrictions and a largely controlled system with very low community spreading, well below the epidemiological threshold. Hence in this initial phase no import had to be added to explain the ongoing dynamics.

After the lifting of the lockdown measures in summer 2020 an increase of severe cases was observed, but then leveling off to relatively stable values, while the increased testing capacities led to a strong increase of positive PCR tested cases, however mostly cases with mild symptoms or even asymptomatic. Recently, levels of around 80% of asymptomatic cases have been reported among positive tested [19]. Isolated outbreaks were increasingly detected. In order to adjust our models to this new scenario given by the data at hand, a new level of import, from the time of releasing the lockdown, was included, and also a changing detection rate of mild or asymptomatic cases were investigated. A dynamical behaviour analogously to what we observed analytically in the simplest SIR-type models with import close to the critical threshold, but still subcritically, was found. Further we assume that import starts mainly by asymptomatic infection, since symptomatic and severe cases would be restricted from mobility or restrict themselves easier than unnoticed infections.

Below we describe briefly the updated models and their dynamical behaviour, comparing with data up to end of October 2020, until then no real signs of a new exponential phase of epidemiological growth were predicted for the Basque Country and so far not observed. The dynamics follow pretty closely the pattern which would be expected from the simple models of SIR-type under import with sub-threshold community spreading, including the large stochastic fluctuations of a system close to criticality. A more detailed analysis of such rather large and complex systems as the SHARUCD-type models can only be performed in due time and will be reported as results to come in. Note, however, that the qualitative description given here describes the epidemiological system much better than any model with a second exponential phases, i.e. adjusting the community spreading with every new stochastic fluctuation.

### 3.1 Model update of SHARUCD to include analysis of isolated outbreaks after lockdown lifting

The model to analyze isolated outbreaks contains now import to asymptomatic infection after lifting of lockdowns and increased detection of asymptomatic due to intensified contact tracing

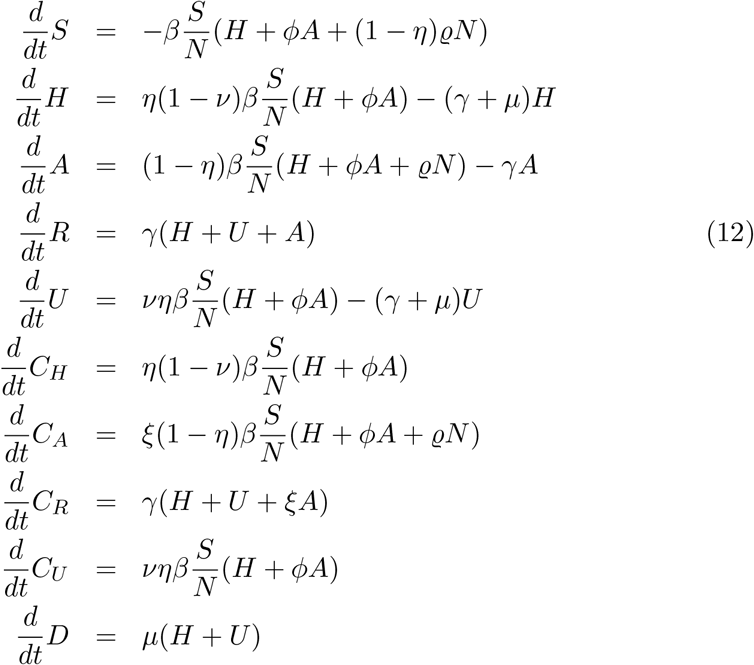

with time dependent infection rate *β*, and now also increased import *ϱ* and increased detection of asymptomatic cases *ξ*.

The import is increased from zero initially *ϱ*_0_ = 0 to a level *ϱ*_1_ > 0 with sigmoidals *σ*_+_ and *σ*_*−*_ as previously the infection rate *β* [1]. We have increasing sigmoidal *σ*_+_(*x*) ≔ 1*/*(1 + *e*^*−x*^) and decreasing sigmoidal *σ*_*−*_(*x*) ≔ 1*/*(1 + *e*^*x*^).

Due to changing PCR-testing strategies, from previously only testing severe cases and now mainly contact tracing finding many more asymptomatic cases, the detection ratio of asymptomatic was increased, i.e. the parameter *ξ* similarly to the increased import *ϱ*. Hence we have *β*(*t*) as described earlier, and now also *ϱ*(*t*) and *ξ*(*t*) as time dependent parameters, changing once to a higher level when the lockdown was lifted in summer 2020.

Critical fluctuations explain the at the moment mainly observed isolated outbreaks, since community spreading, measured by the infection rate *β* is still slightly below the threshold value *β*_2_ < *β*_*c*_, but imported cases, measured by *ϱ*, are occasionally sparking larger outbreaks of mainly asymptomatic or mild, and occasionally also via these producing few sever case, since *β*_2_ ≈ *β*_*c*_. This explains the small numbers of severe cases at the moment, but a strong increase in asymptomatic or mildly infected due to the increased success of systematic contact tracing.

### 3.2 Real data supporting theory: subcritical regime to explain COVID-19 spreading in the Basque Country

Rather than a new exponential growth phase, the recent data from the Basque Country show small increases in sever cases to its leveling off, from August 2020 to at least end of October 2020, portrayed already as a “second wave”. These data described well the predicted spreading observed in a subcritical regime with finite import factor included in the model after the first lockdown was lifted. As for now, the SHARUCD modeling framework continues to predict stationarity for all variables. It is, however, still early to know to which lower level the disease transmission will develop, as community transmission is even lower due to the second lockdown. Nevertheless, the community spreading could be expected to increase significantly at the onset of the respiratory disease season in winter.

Fig. 6 shows the comparison between the stochastic SHARUCD model realizations (mean are plotted as light blue line) and empirical data (cumulative cases are shown as black dots), described earlier in [1], where all basic parameters are kept from the calibration during the initial phase of the epidemic. We also show the daily incidences of positive PCR tested persons in Fig. 7 a), the hospital admissions in b), the ICU admissions in c) and the deceased cases in d). Empirical data are plotted as black line, mean are plotted as light blue line and the 95% confidence intervals (CI) are plotted in light purple (shadow). The observed dynamics are described by severe cases of hospitalized and ICU admissions as well as the deceased cases increasing during the initial exponential growth in March 2020, decreasing significantly during the first lockdown, i.e. the controlled phase from April to the end of July 2020. From the beginning of August onwards, after the complete lockdown lifting, a slight increase in cases are observed, leveling off to stationarity with constant number of cases, with large fluctuations around the mean. However, the number of cases are in the range of the predicted 95 % CI of the stochastic SHARUCD model, which are themselves subject to fluctuations since the CI are calculated directly from the ensembles of stochastic realizations.

**Figure 6:**
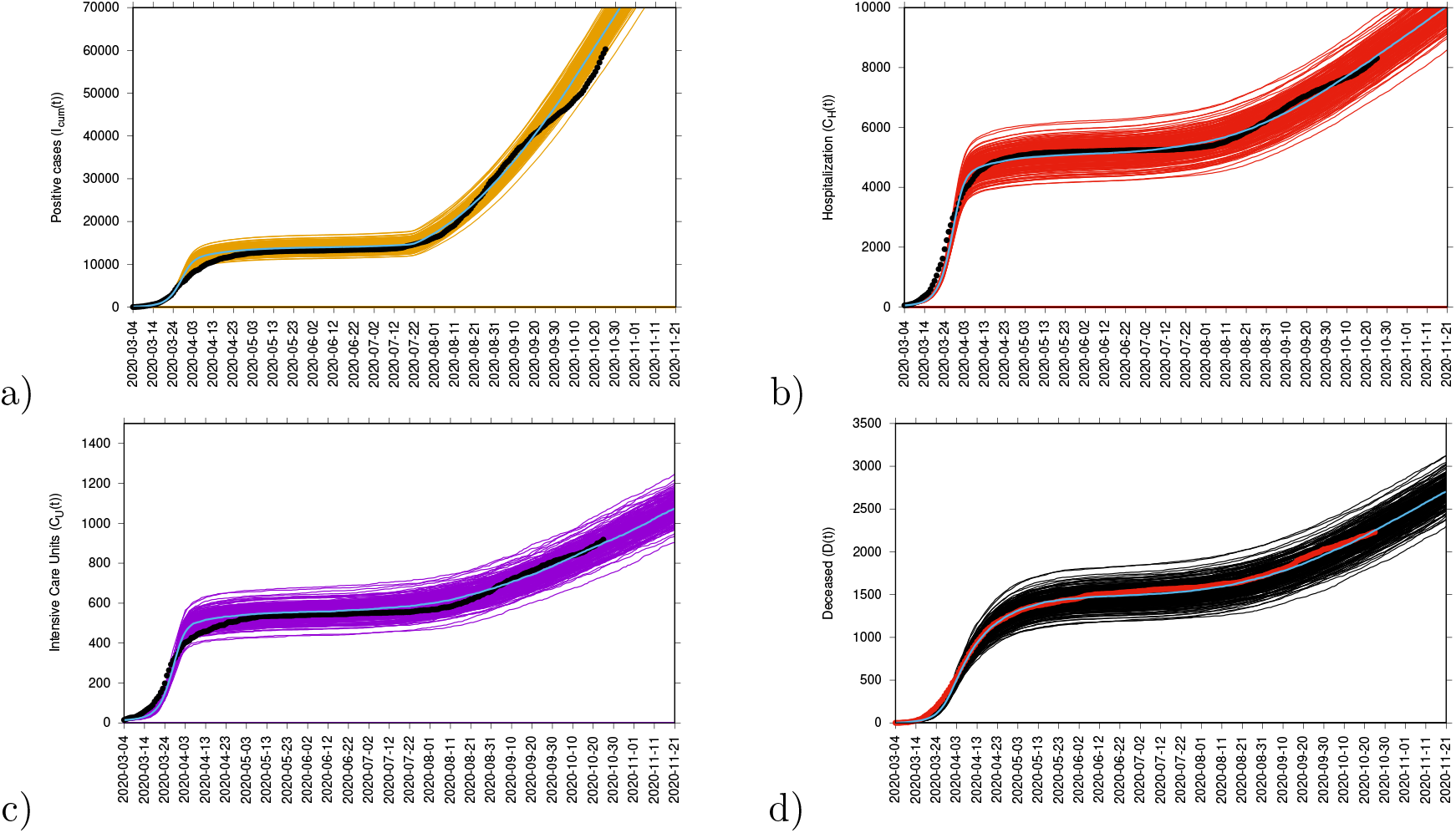
Cumulative cases updated for a) positive tested, b) hospitalized, c) ICU admitted and d) deceased cases, until end of October 2020.

**Figure 7:**
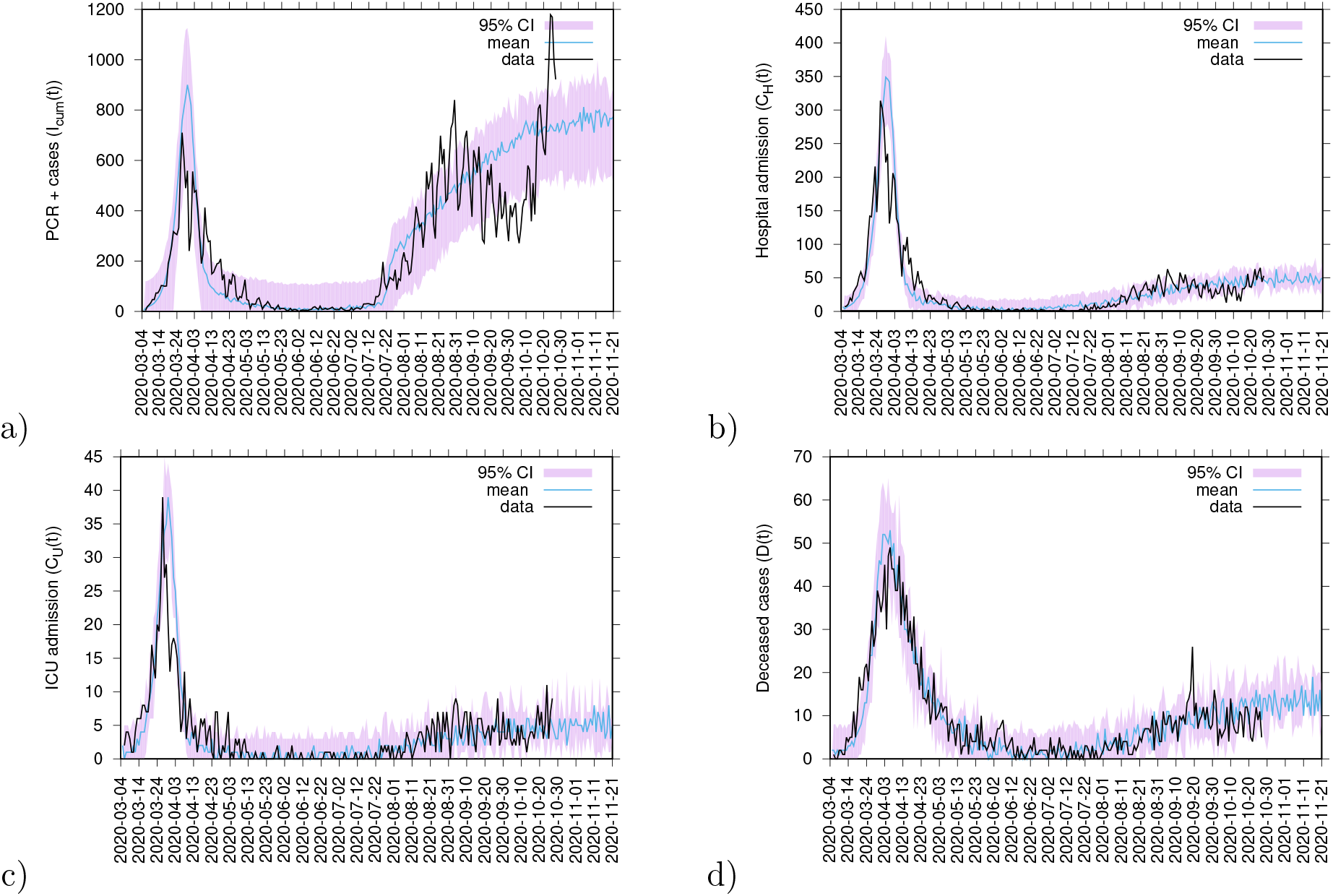
Incidences updated for a) positive tested, b) hospitalized, c) ICU admitted and d) deceased cases, until end of October 2020. Here we see the initial increase to a second phase, of disease spreading from late July and August 2020 on, so called “second wave”, but then the leveling off to a stochastic fluctuating stationary behaviour.

These qualitative behaviour of a slight increase followed by settling into a fluctuating stationary state is observed and hold for a long time, from August to end of October 2020, when the new lockdown was implemented, in very good agreement with the critical behaviour in the simple SIS and SIR systems under the influence of a conjugate field of import. The empirical data support this theory, preliminarily analyzed in Section 2 of this manuscript. The term “import” is used here as described in mathematical terms, any transition of susceptibles without any detected contact with an infected from the study-population, a quite broad definition, but well distinguished now in the new phase after lifting of the lockdown from any large supercritical increase of community spreading.

A final word has to be given to the rather untypical behaviour of the positive detected cases, shown in Fig. 7 a). We could only describe the empirical data by increasing vastly the detection rate of asymptomatic, which does not affect at all the other variables in b) to d). We might have even underestimated the initial level of asymptomatic cases in spring 2020, when only severe cases were tested via PCR due to the limited testing capacity at that time. The situation has changed significantly and with many asymptomatic cases detected by increased community testing, screening campaigns and a better contact tracing strategy. Like that, to describe quantitatively the behaviour of detected positive cases the SHARUCD model must be adjusted to changing testing capacities rather than the disease dynamics itself, i.e, by increasing the community transmission, which is reflected in the severe cases dynamics. The predicted qualitative behaviour is consistent with the theory with positivity rates kept stable, even with an increased testing capacity.

The situation is feared to change again during the onset of the respiratory disease season, when community spreading might increase naturally without any behavioural control changes, such that community spreading could push the system above threshold into a real new exponential phase, visible in semilogarithmic plots, as we showed for the first exponential phase in [1]. Hence, a “real second wave” as observable in SIR systems with recurrent outbreaks could describe the disease spreading behaviour in a supercritical community transmission regime.

As the Basque Country faces a second lockdown phase since October 27, 2020, this danger has rather decreased, and the system might be controlled just below the epidemiological threshold for a longer time, with large fluctuations to be expected. That is because the second lockdown and the control measures to avoid COVID-19 have not only decreased the community transmission even more, but also affected the transmission of other respiratory diseases that might occur as a smaller outbreak than observed in years prior to the COVID-19 pandemic.

From our analysis we conclude that a mild control to keep the system sufficiently below threshold to avoid large critical fluctuations, but allowing a smooth economic recovery and better planing of necessary long term maintenance activities in many areas of life is the way to decrease disease speeding impact on our society, until reaching the herd immunity threshold by vaccination strategies [14, 15]. Our analysis is based on the observation from the empirical data and our models that natural herd immunity due to burn out of susceptibles is not yet close to be reached, though we have since the beginning taken large proportions of asymptomatic infected into account aside the severe cases. So the assumption of abundance of susceptibles is still giving good results, as shown here. And research questions remain as to which extend classical vaccines against COVID-19 [14] with comparable efficacies as other vaccines developed over decades for different diseases [16, 17, 18] will be needed for effective control or new vaccine lines can keep their promising high efficacies even in continued evaluation over the next year.

## Data Availability

Epidemiological data used in this study are provided by the Basque Health Department and the Basque Health Service (Osakidetza), continually collected with specific inclusion and exclusion criteria.

https://www.ecdc.europa.eu/en/covid-19/data

https://www.euskadi.eus/boletin-de-datos-sobre-la-evolucion-del-coronavirus/web01-a2korona/es/

https://www.mscbs.gob.es/profesionales/saludPublica/ccayes/alertasActual/nCov/

## 4 Acknowledgments

Data were provided by the Basque Health Department and the Basque Health Service (Osakidetza). We thank the huge efforts of the whole COVID-19 Basque Modelling Task Force (BMTF), specially to Eduardo Millán for collecting and preparing the data sets used in this study. We thank Adolfo Morais Ezquerro, Vice Minister of Universities and Research of the Basque Government for starting the BMTF initiative and for the fruitful discussions. M. A. has received funding from the European Unions Horizon 2020 research and innovation programme under the Marie Skłodowska-Curie grant agreement No 792494. This research is also supported by the Basque Government through the BERC 2018-2021 program and by Spanish Ministry of Sciences, Innovation and Universities: BCAM Severo Ochoa accreditation SEV-2017-0718. N.St. thanks the Mathematics Department of Trento University for the opportunity and support as guest lecturer in biomathematics, during which part of the present work has been conceived and started to be developed.

## Appendices

### A Analytical solutions for the SIS model with import *ϱ*

In this appendix we give the analytical time dependent solution of the SIS system with import in its mean field approximation, which then can be analyzed further in terms of scaling behaviour around the critical threshold in the next appendices. This system has a paradigmatic status for such investigations of scaling around criticality, being an important member of the universality class of directed percolation, whereas the SIR system in its spreading regime into an abundant environment of susceptibles is a member of the dynamic universality class, which has attracted somehow less attention, but seems more relevant in real epidemiological settings as discussed here in the main text.

The SIS-system model with import *İ* = (*β/N*)(*N* − *I*)(*I* + *ϱN*) − *γI* can be given in reduced form as

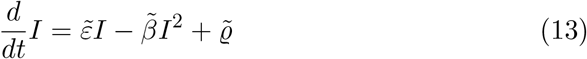

with 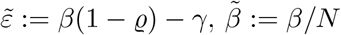 and 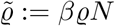.

Here we explore the possibility of an analytical solution analogously to the basic SIS model without import, as long as possible to understand the structure of the analytic solution. The solution of the SIS model with vanishing import *ϱ* = 0 is given by

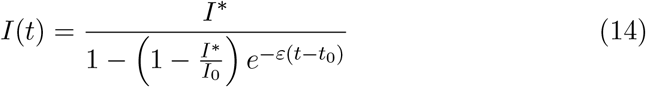

with *I*^*\**^ ≔ (1 − *γ/β*)*N, ε* ≔ *β* − *γ* and initial condition *I*_0_ ≔ *I*(*t*_0_).

An analogous calculation gives for the case of non-vanishing import the result of

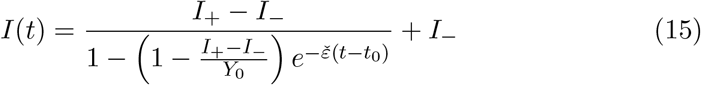

with the shifted constants, as compared to the vanishing import case, 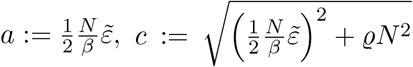, giving *I*_+_ ≔ *a* + *c* and *I*_−_ ≔ *a* − *c*, further *Y*_0_ ≔ −*I*(*t*_0_) − *I*_*−*_ and 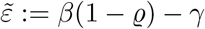 and for the exponential rate 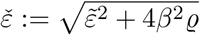 with solutions given numerically in Fig. 8. Equivalent solutions and further considerations on the stochastic version of the SIS system with import have been reported in [11] with e.g. highly non-Gaussian stationary distributions close to the epidemic threshold of extinction.

**Figure 8:**
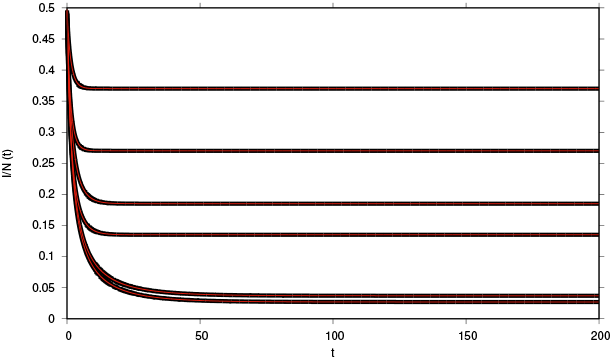
Analytical solution I(t) in red, compared with the numerical solution via integration of the dynamics 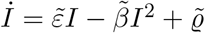 in black.

### B Unifying framework for criticality in epidemiological systems

When it comes to threshold behaviour in epidemiological systems it has been for a long time established that the so-called “general epidemic process” (GEP), describing the infection of susceptible individuals and their subsequent recovery (SIR), falls into the universality class of dynamical percolation with some of the critical exponents agreeing with ordinary bond percolation [7], while for the simplest epidemiological model, with only susceptibles being infected and infected becoming directly susceptible again without recovering (SIS), falls into the universality class of directed percolation [10].

The additional effect of import in this simplest SIS case has more recently been investigated in detail as “conjugated field”, like in equilibrium phase transitions an external magnetic field influencing the critical behaviour of spin systems [9].

Of major importance in the spreading of epidemiological systems is the behaviour in two dimensional space or in case of long ranging contacts or homogeneous mixing the mean field behaviour. We will consider the critical exponents’ values in these two cases. Recently, for the SIR-type models the interpolating behaviour between mean field behaviour and purely local spreading in two dimensions due to super-diffusive spreading has been investigated by Grassberger [8] in great detail.

We will first investigate the SIS model with import, for which a complete analytic solution can be given in mean field approximation, both for the static stationary behaviour as well as for the dynamic time dependent solution. This establishes the critical exponents and universal scaling functions in its mean field approximation.

Then we investigate the SIR model with import in the same way, where the stationary state solution can be calculated as well, when we assume waning immunity in addition to the GEP model, which has by default no transition from recovered to susceptibles again, hence no nontrivial stationary state. The inclusion of waning immunity allows to compare directly the critical behaviour of SIS and SIR models with import for very long times *t* → ∞, having the same stationary exponents in mean field approximation.

In the limit of vanishing waning immunity *α* we can analyze for large population size *N* and hence susceptibles in abundance *S/N* ≈ 1 the critical behaviour, which is again the same in the SIS and the SIR case, and holds for epidemiological systems in their initial phase of exponential growth supercritically or exponential decline subcritically, as well for vanishing import as for import larger than zero. This is the scenario described in the main text.

#### B.1 SIS and SIR epidemic models and their stationary behaviour

The full SIR model with import is given by

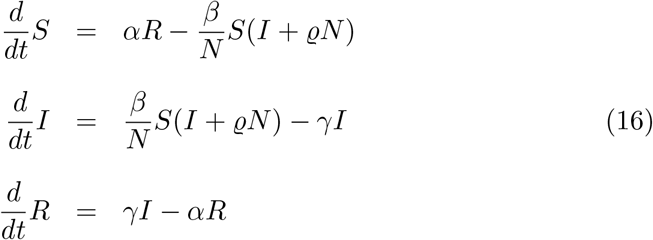

from which the SIS model can be derived by neglecting the recovered class or assuming the transition from *R* to *S* as infinite *α* → ∞ and with conserved population size *N* = *S*(*t*) + *I*(*t*), hence *S* = *N* − *I*, given by

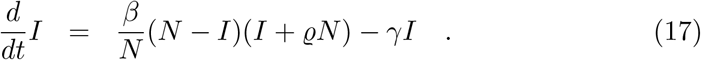

Further the general epidemic process (GEP) follows from the SIR model via the limits of vanishing waning immunity *α* = 0 and vanishing import *ϱ* = 0 as

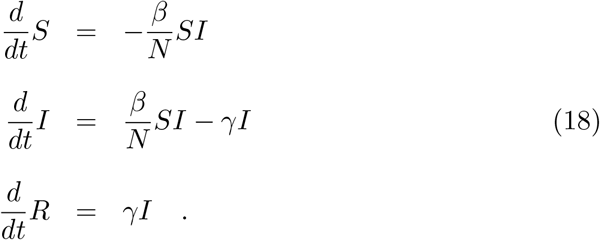

And finally the case of infinitely many susceptibles available in an infinite population of size *N*, hence the case of *S/N* ≈ 1, now including again import *ϱ* > 0, is in mean field approximation simply given by

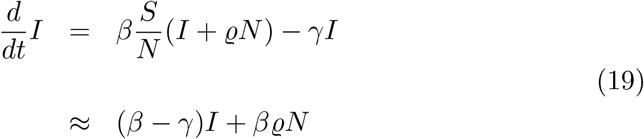

as a basic model for any spreading of infection in a susceptible population with no limiting availability of resources, here the susceptibles, and including import.

For the SIS model with import we have the stationary state

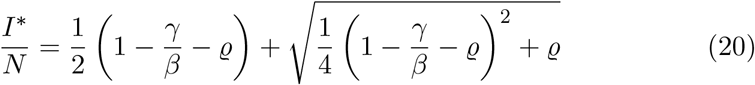

and for the SIR model with import and waning immunity analogously

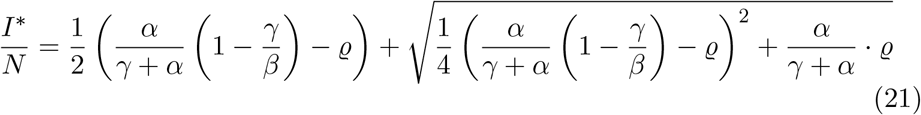

In both cases we have for vanishing import *ϱ* = 0 and in the case of the SIR for positive waning immunity *α* > 0 the scaling

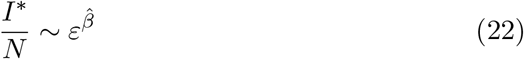

with *ε* ≔ *β* − *β*_*c*_. In mean field approximation we have *β*_*c*_ = *γ* an for finite import *ϱ* > 0 and vanishing *ε* = 0

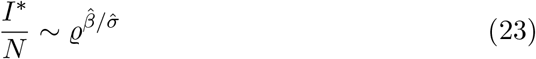

with critical exponents 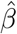 and 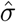. To distinguish the epidemiological parameters *β, γ* etc. from the also used Greek letters in the literature of non-equilibrium phase transitions, we use 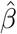 etc. for the critical exponents. The notation of exponents otherwise is close to the ones defined in [9], which are often close to other references, but not always, in part depending how close the notations are to earlier established equilibrium phase transition notations, as e.g. sketched again in [9] or earlier percolation literature and its adaptation [10, 7], and developed from there [8].

The above given scaling of the power laws hold for the SIS system, and also for the SIR system when we have non-vanishing waning immunity *α* > 0, since only then a non-trivial stationary state is possible.

#### B.2 Dynamical behaviour and scaling

For the SIS-system we have a complete time-dependent analytical solution, and hence can also investigate rigorously the scaling with time going to infinity *t* → ∞ along the analysis with the other static parameters *ε* → 0 and import *ϱ* → 0.

The basic reduced model of the SIS-system with import we have from the original model *İ* = (*β/N*)(*N* − *I*)(*I* + *ϱN*) − *γI* as given in Eq. (3) the dynamic equation

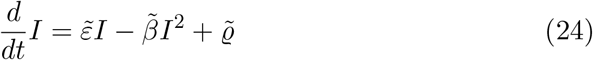

with 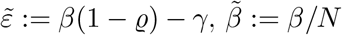 and 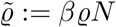. The time dependent solution is given in Appendix A.

This establishes the scaling behaviour around criticality for the density of infected 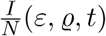 as scaling ansatz

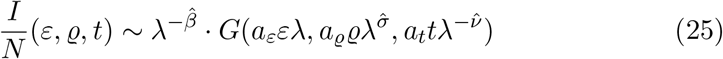

with critical exponents 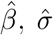 and 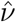, where here the time exponent sometimes called *ν*_*‖*_ is ment, and a common scaling quantity *λ* and amplitudes *a* for the respective parameters *ε, ϱ* and *t*, and a universal function *G*, see [9] for more details.

Fixing *λ* e.g. via the condition *a*_*ε*_*ελ* ≔ 1, hence *λ* = (*a*_*ε*_*ε*)^*−*1^ gives the qualitative asymptotic behaviour (now not taking the amplitudes into account any more, since we deal with individual models where *G* can include the explicit constants) as

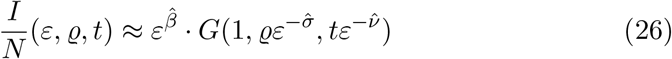

modulo amplitudes *a*.

Respectively fixing *λ* via 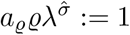 gives

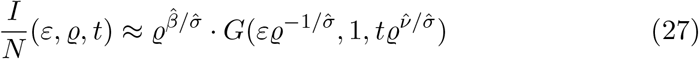

and for scaling with time *t*

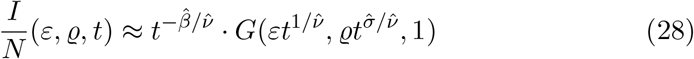

or at criticality and vanishing import, hence *G*(0, 0, 1) = *const*.,

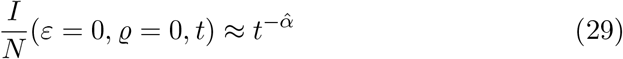

hence as commonly used exponent 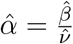.

From inspection of the time dependent solution for *I/N* we can read off the critical exponents in mean field approximation

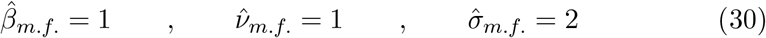

and hence also with 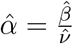

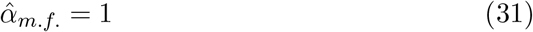

see e.g. [9].

For two-dimensional spatially explicit models in this universality class of directed percolation we have the exponents

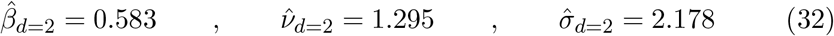

and hence 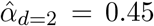, hence for spatially extended versions of SIS models with import as well [9].

For the SIR model we do not have any time dependent analytically closed expression even in mean field approximation at hand, but the stationary state scaling already indicates that the above mentioned critical behaviour also should hold here, as long as waning immunity is non-vanishing and time dependence ultimately is determined close to this stationary state behaviour.

However, for the initial spreading in very large systems and not yet relevant waning immunity, i.e. mathematically *α* = 0, and system size *N* going to infinity and hence infinitely many susceptibles available, there has been suggested another scaling in so-called spreading experiments for the general epidemic process GEP, see [7]. A simple mean field analysis we sketch now in the next subsection.

#### B.3 SIR spreading in simplified mean field approximation, GEP limit

The SIR epidemic model as well as any other with susceptibles in abundance, hence *S/N* ≈ 1, is given by

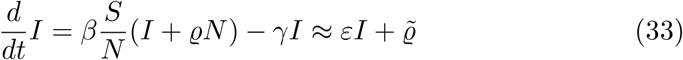

with *ε* = *β* − *β*_*c*_ = *β* − *γ* and 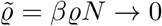, hence in the limit of *N* → ∞, but *ϱ* → 0 faster (which can be to some extend cast into finite size scaling [9]).

We have the time dependent solution

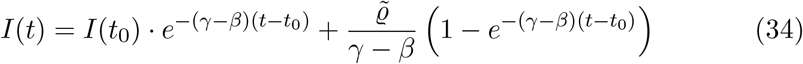

and at criticality *ε* = 0 we have the solution

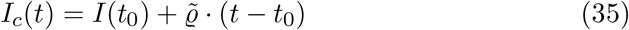

hence at criticality and vanishing import *ϱ* = 0 the dynamical behavior

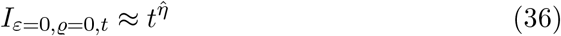

with a new critical exponent 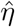, see [8] with value in mean field approximation

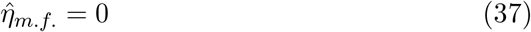

(which is identified with 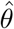 in [9]) and in two spatial dimensions

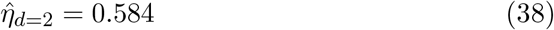

see [8], including interpolating behaviour for super-diffusive spreading from mean field behaviour to purely local spreading in two dimensions, GEP with ordinary percolation identifiable [7]

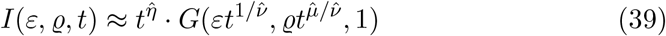

hence mean field exponents 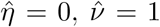 and 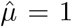 as can be seen from the time dependent solution in mean field approximation (with 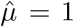 unequal 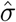 from the SIS case).

### C Data collapse plots for scaling analysis

Data collapse plots confirm the scaling behaviour as described above and characterize the self-similarity close to critical thresholds. We describe in the following only the principles as they can already be seen from the mean field analysis. But all holds for spatial and network systems and can experimentally be investigated without analytic solutions at hand, see e.g. various such plots in [9, 7] and many other research publications.

#### C.1 Static scaling in SIS with import for stationary solution

For the static behaviour of the SIS model with import we have for the density the explicit solution, see Eq. (20), given by

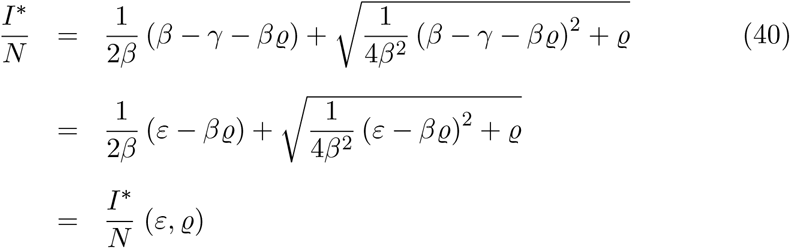

and close to criticality *ε* = 0 and *ϱ* = 0

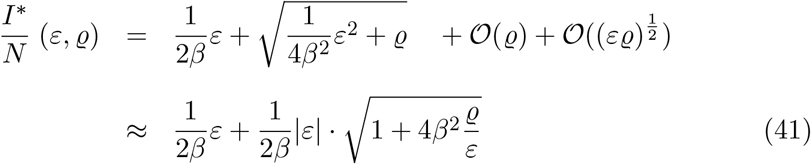

From the second line in Eq. (41) we see that for *ϱ* = 0 we have the power law

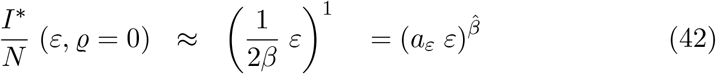

and for *ε* = 0 the power law

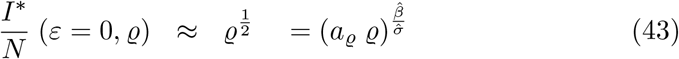

and from the scaling ansatz

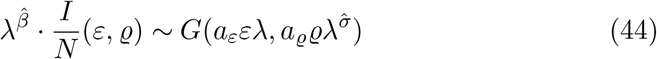

with *a*_*ϱ*_*ϱλ* ≔ 1, hence fixing *λ* as 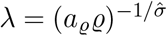 we obtain for the data collapse plot

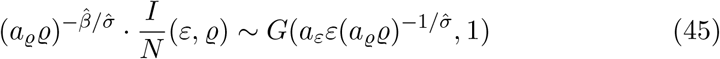

with the universal scaling function in mean field approximation

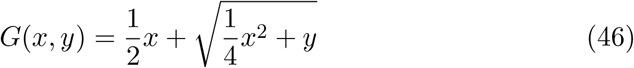

with the factor of 1*/*2 to normalize to *G*(0, 1) = 1.

Then we plot Eq. (45) for 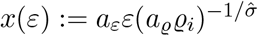 for various values of *ϱ*_*i*_ and varying continuously *ε* the quantity 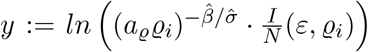, using the logarithm only to obtain reasonable numbers. The sequence of imports follows *ϱ*_*i*_ → 0. For comparison we also plot

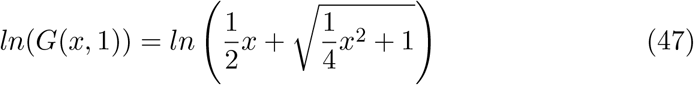

to which the data collaps converges for small *ε* and small *ϱ*_*i*_.

#### C.2 Dynamical scaling with time of the SIS model with import

For *ϱ* = 0 we have the scaling of *t* · *I/N* (*ε, ϱ* = 0, *t*) = *F* (*εt*) as given in the following graphics.

We use here for simplicity the dynamical equation for the densities of infected *I/N* (*t*) given by

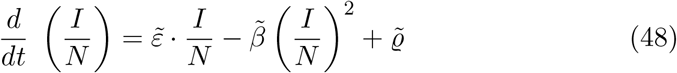

and for vanishing import *ϱ* = 0 the values 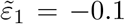, and −0.05, −0.01 and then positive 0.01, 0.05 and 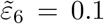. Initially we have half the population infected, *I*(*t*_0_)*/N* = 0.5.

This means, the values for *β*_1_ are *β*_1_ = 0.9 · *γ*, then 0.95, 0.99, 1.01, 1.05 and 1.1 in the case of vanishing import *ϱ* = 0, see Fig. 11, when using *γ* = 1.

**Figure 9:**
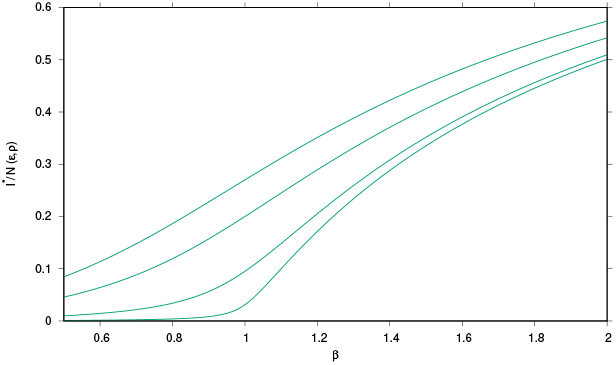
Stationary state density I^*^/N varying with β for ϱ_1_ = 0.1, ϱ_2_ = 0.05, ϱ_3_ = 0.01 and ϱ_4_ = 0.001.

**Figure 10:**
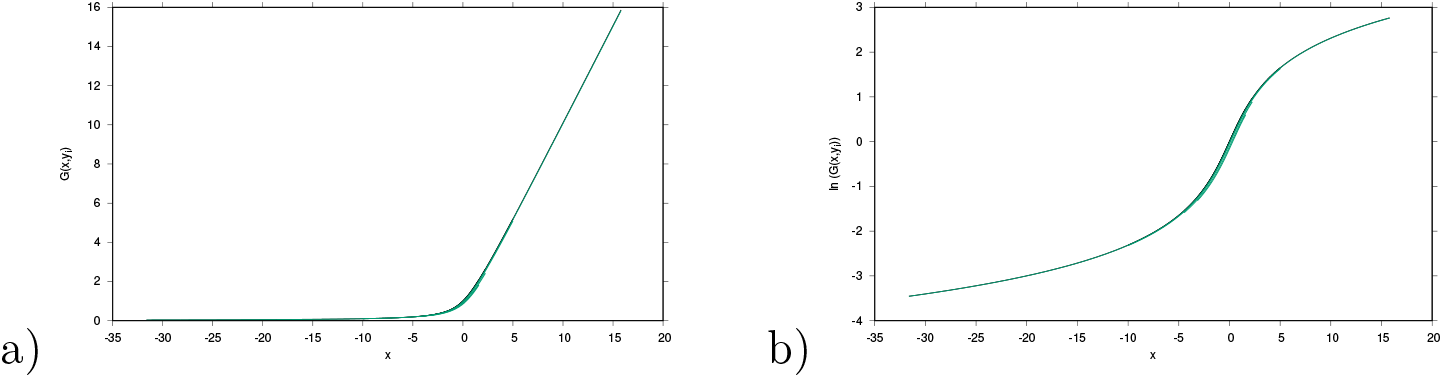
a) Scaling function 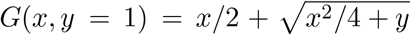, and data collpapse of the stationary state density I^*^/N for ϱ_1_ = 0.1, ϱ_2_ = 0.05, ϱ_3_ = 0.01 and ϱ_4_ = 0.001, and ε varied from ε = −0.5 to ε = 1, hence β = γ + ε varies from β = 0.5 to β = 2. b) Same for ln(G). In both cases we use 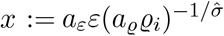 and 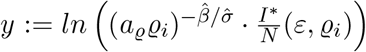 with 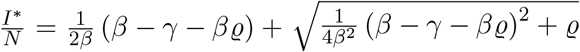 and a_ε_ = 1/β and a_ϱ_ = 1, 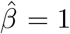 and 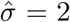. Here we simply use γ = 1, hence time scales with inverse recovery rate.

**Figure 11:**
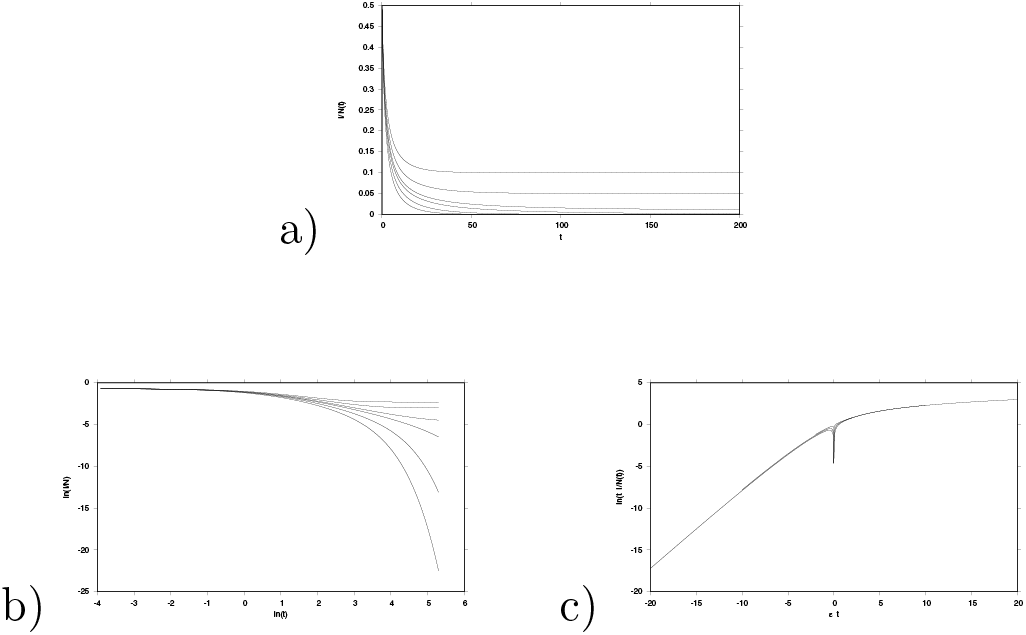
For vanishing import *ϱ* = 0 we have the time dependence a) in natural scale b) in double logarithmic scale and in c) as scaling of data collapse.

For vanishing distance to criticality along *ε* = *β* − *γ*, we have *ϱ*_1_ = 0.2, then 0.1, 0.05, 0.01, 0.005 and 0.001., see Fig. 12. Hence for the scaling with *ϱ* and vanishing distance to criticality *ε* = 0 we obtain similar data collapse plots, as well as for their combinations, i.e. non-vanishing import and non-vanishing distance to criticality. Likewise the spreading in the GEP can be analyzed analogously, giving the universal function and the respective universal critical exponents, as used in the main text.

**Figure 12:**
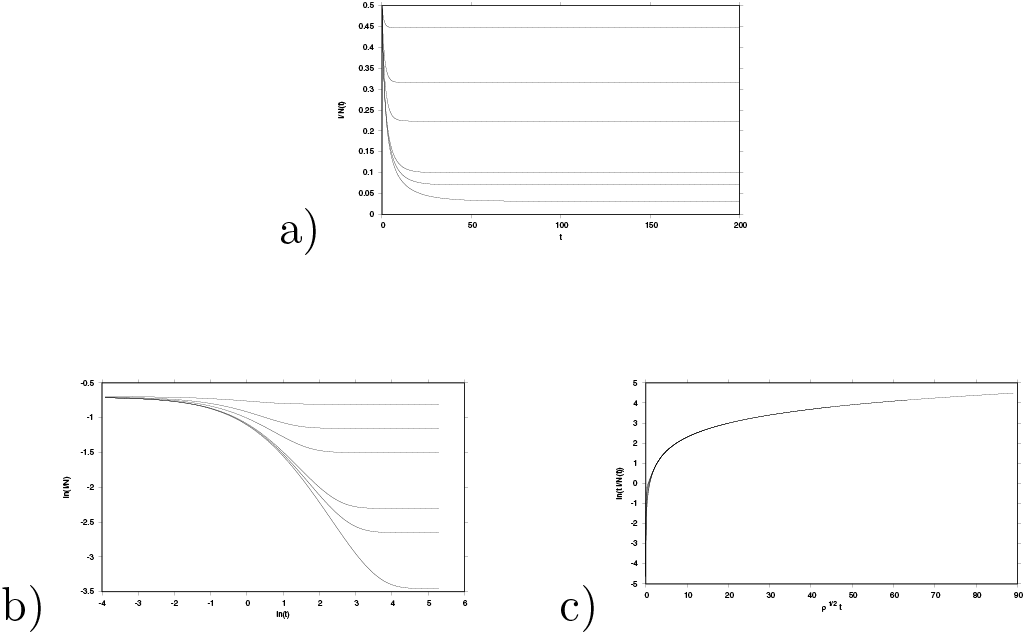
For vanishing distance to criticality *ε* = 0 we have the time dependence a) in natural scale b) in double logarithmic scale and in c) as scaling of data collapse.

#### C.3 Spanish data and other European countries

In Figure 13 we plot histograms of weekly new positive cases (yellow), hospital admissions (red), ICU admissions (purple), and deaths (black) associated to COVID-19 for Spain, the Netherlands, Belgium, and France. Data were collected from the website of the European Centre for Disease Prevention and Control (ECDC) [22]. Hospital and ICU admissions were reported per 100,000 people and were here rescaled according to the total population of each corresponding country (as of the end of 2019) as reported in the ECDC data spreadsheets. The first epidemiological week of 2020 for which data was available varied between countries (hospitalizations: week 5 Spain, week 11 Belgium and France, week 7 Netherlands; ICU admissions: week 5 Spain, week 22 Belgium, week 9 Netherlands, week 11 France) while the last considered week in this work for all countries was week 44 (ending on October 31st 2020).

**Figure 13:**
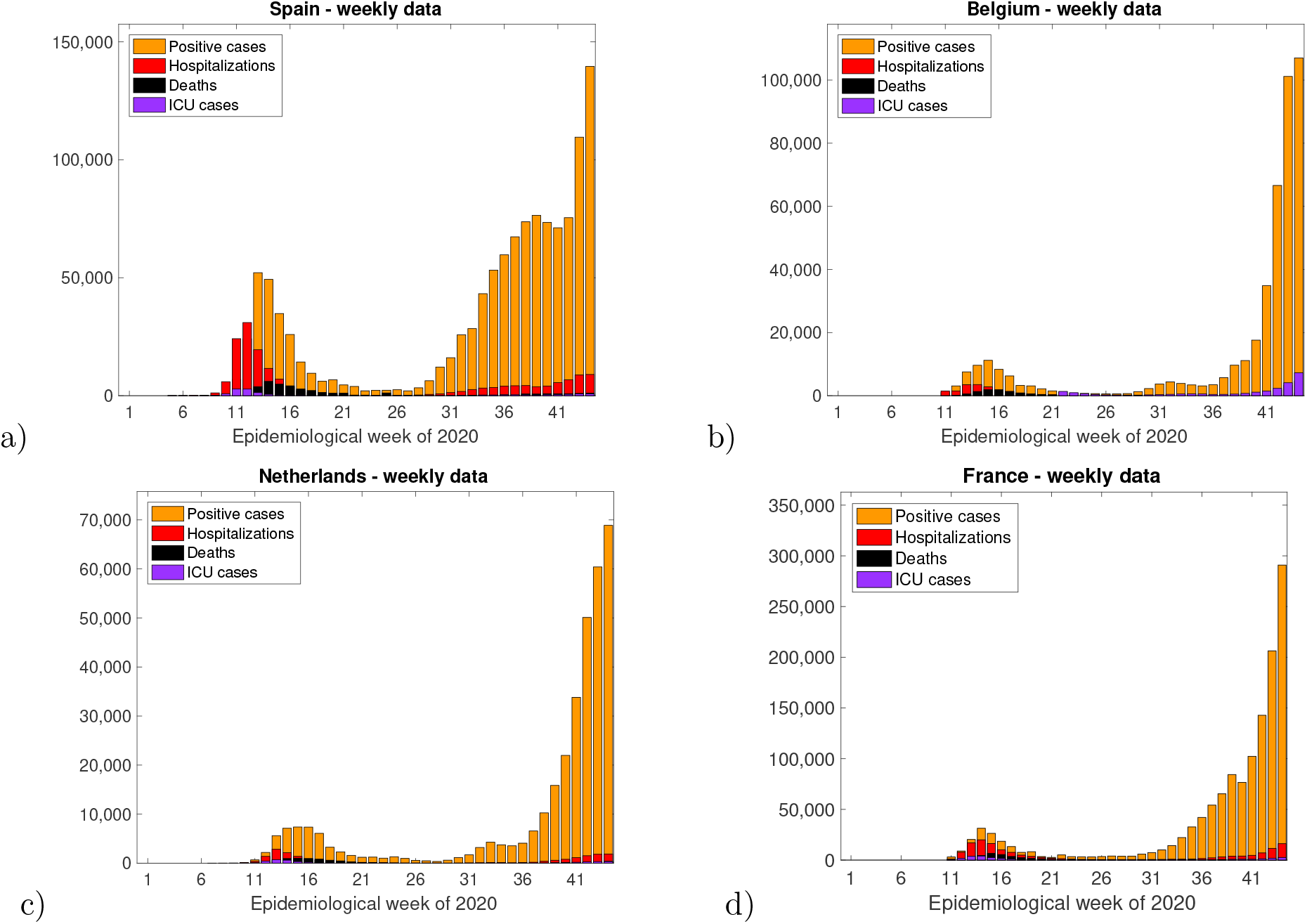
Weekly positive cases, hospitalizations, ICU cases, and deaths in a) Spain, b) Belgium, c) the Netherlands, and d) France until the end of epidemiological week 44 (31st of October) of 2020.

Figure 14 shows histograms of weekly positive cases (yellow) and PCR tests performed (light blue) for the same countries. To better understand changes in the number of positive cases versus testing capacity over time, we superimpose the percentage (red lines) of positive tests over the total number of tests performed obtained from the histogram data for each considered week. Once again the first epidemiological week of 2020 for which data on testing was available varied between countries (week 18 Spain, week 9 Belgium and France, week 11 Netherlands) while the last considered week for all countries was week 44.

**Figure 14:**
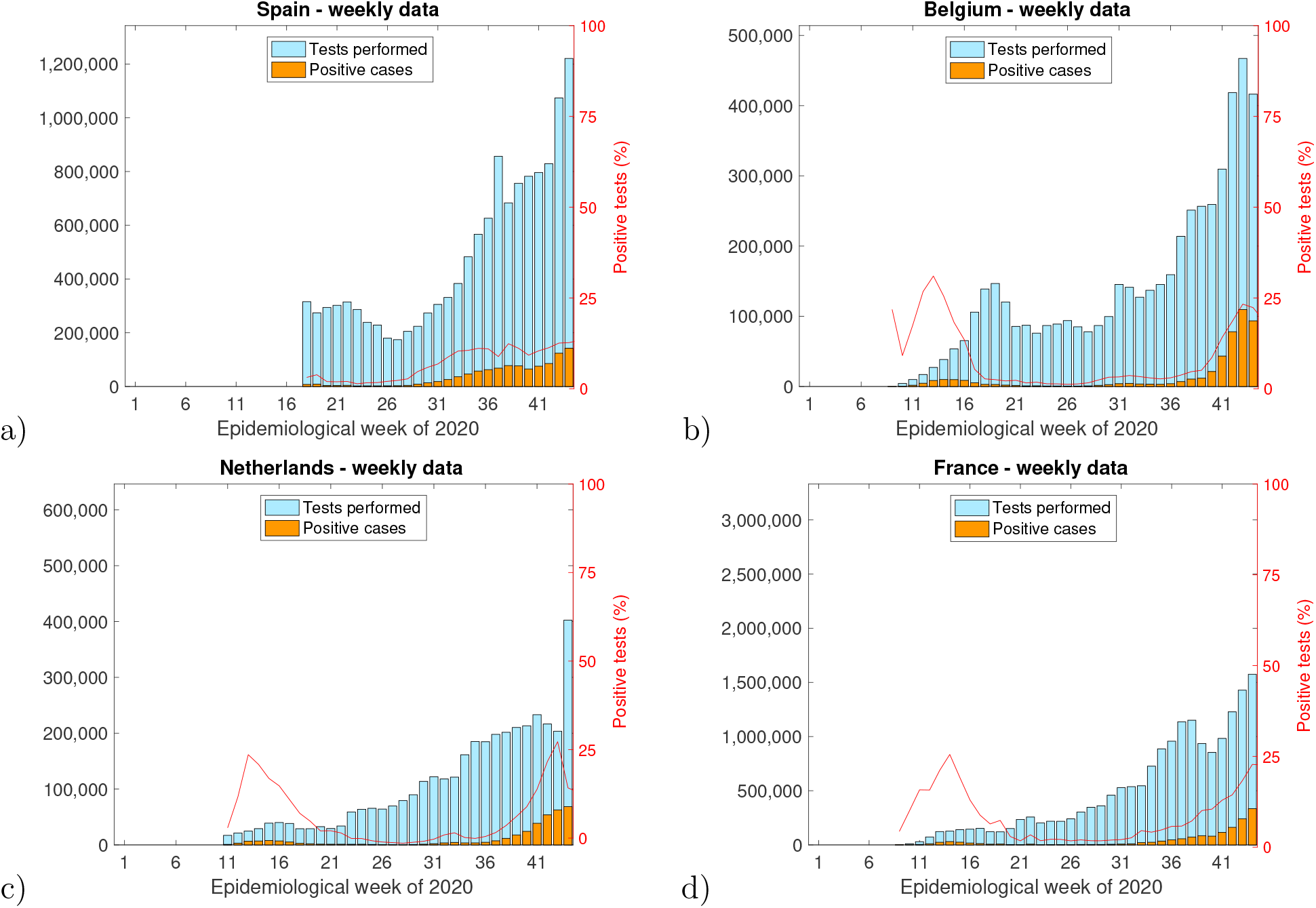
Weekly positive cases and number of PCR tests performed in a) Spain, b) Belgium, c) the Netherlands, and d) France until the end of epidemiological week 44 (31st of October) of 2020. The percentage of positive tests (red lines) was computed by dividing the number of weekly positive cases by the number of total PCR tests performed in the same week multiplied by 100.

Finally, in Figure 15 we show the evolution throughout the year 2020 of the case-fatality ratio (CFR) for the considered countries. The value of the CFR was obtained by dividing cumulative deaths by cumulative positive cases and multiplying the result by 100 for CFR given in per cent. Cumulative data were computed from daily data on positive cases and deaths once again obtained by the ECDC COVID-19 dataset webpage for all countries [22].

**Figure 15:**
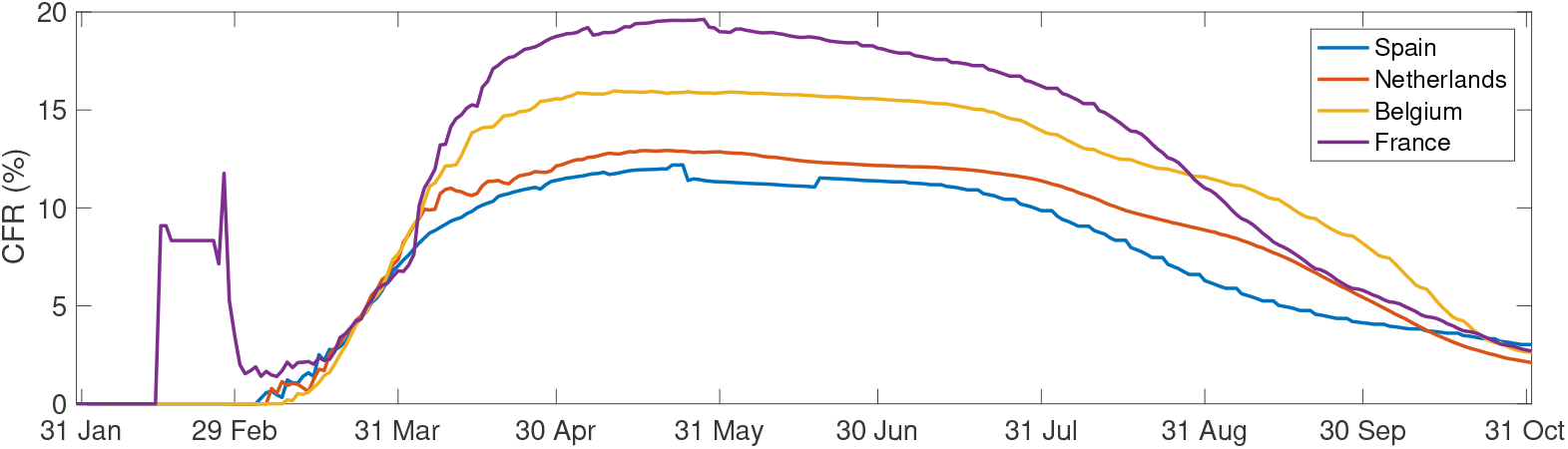
Evolution of the case-fatality ratio for Spain, the Netherlands, Belgium, and France until the end of October 2020.

In conclusion we observe that though the detected infected cases increased in the countries investigated here in the second half of the year 2020 to a large amount, but severe cases did not keep the same increase, hence increased testing capacities detected mainly mild or asymptomatic cases which previously in the first half of the year 2020 would have gone unnoticed. Large increases of detecting mild or asymptomatic cases consequently also decreased the case fatality ratios in the various countries, eventually converging to similar magnitudes reflecting biological universality of this measure, whereas the large differences in the various countries since the beginning of the epidemic seem to have reflected differences is detection of mild cases and initial fluctuations of cases in various outbreaks in the different countries.

